# Greater pericardial and visceral adiposity increase risk of hospitalization or death from SARS-CoV-2 infection in community dwelling adults: A C4R Study

**DOI:** 10.1101/2024.10.02.24314721

**Authors:** Nadine Al-Naamani, Pallavi Balte, Michael G.S. Shashaty, Asma Sharaf, James G. Terry, John Jeffrey Carr, Matthew Allison, Cheryl A.M. Anderson, Surya P. Bhatt, Michelle T. Long, Jason D. Christie, Elizabeth Oelsner, Michaela R Anderson

## Abstract

**Importance:** Higher body mass index (BMI) increases risk of respiratory failure and death from SARS-CoV-2 infection. BMI is imprecise and fails to account for adipose tissue distribution.

**Objective:** To determine whether pericardial adipose tissue (PAT), abdominal visceral adipose tissue (VAT), subcutaneous adipose tissue (SAT), or hepatic adipose deposition on pre-pandemic computed tomography (CT) scans associate with increased risk of hospitalization or death from SARS-CoV-2.

**Design:** The Collaborative Cohort of Cohorts for COVID-19 Research (C4R) ascertained SARS-CoV-2 outcomes among participants from 14 US-based cohort studies. This analysis includes participants enrolled in the Jackson Heart, CARDIA, MESA, and Framingham Heart studies.

**Participants:** C4R attempted to enroll all cohort participants who were alive on March 1, 2020 and had not withdrawn consent for cohort participation.

**Exposures:** Adipose depot size measured on research CT scans performed between 2000 and 2011.

**Outcome:** Time from March 1, 2020 to hospitalization or death from SARS-CoV-2 infection ascertained by self-report or active surveillance and confirmed when available, by protocolized record review.

**Results:** There were 8412 participants with at least one CT adipose measure, among whom 184 events occurred over a median (interquartile range) of 547 (338-609) days. Participants were 57% female with median (SD) age of 69.1 (10.4) years. In adjusted models, both higher PAT (per doubling in PAT volume, HR 1.62, 95% CI 1.25-2.09) and higher VAT (per 1-standard deviation, HR 1.41, 95% CI 1.20-1.67) were associated with greater hazards of hospitalization or death from SARS-CoV-2. Associations remained after adjustment for BMI, diabetes, cardiovascular disease, and hypertension. SAT was associated with hospitalization or death from SARS-CoV-2 but not after adjustment for BMI, diabetes, cardiovascular disease or hypertension. Hepatic adipose deposition was not associated with hospitalization or death. Associations did not vary by sex, age, race, cohort, smoking status, diabetes, or BMI.

**Conclusions and Relevance:** Greater CT-measured pericardial and abdominal visceral adipose tissue were associated with increased hazards of hospitalization or death from SARS-CoV-2 independent of BMI or clinical cardiometabolic comorbidities. Further research should investigate use of CT adipose measures to quantify risk of severe disease from viral infections and to investigate mechanisms linking adiposity to lung injury.

**Key Points:** 

**Question:** Is ectopic adipose deposition measured on pre-pandemic CT scan associated with risk of respiratory failure from SARS-CoV-2?

**Findings:** In this retrospective cohort study of 8812 participants enrolled in 4 large cohort studies pre- pandemic, greater pericardial and abdominal visceral adipose tissue were associated with increased risk of severe SARS-CoV-2 infection independent of body mass index, diabetes, hypertension, and cardiovascular disease.

**Meaning:** Pericardial and abdominal visceral adipose tissue are associated with increased risk of hospitalization or death from SARS-CoV2 independent of existing measures of obesity; use of these measures could improve our risk assessments.

## Introduction

Obesity, defined by body mass index (BMI), is associated with an increased risk of testing positive for severe acute respiratory syndrome coronavirus-2 (SARS-CoV-2),^1,2^ as well as for adverse COVID-19 outcomes including severe disease,^3–8^ intensive care,^9^ and death.^8,10,11^ Obesity is also a known risk factor for acute lung injury in other settings.^12–18^ Nonetheless, the mechanisms linking obesity to lung injury are incompletely understood.

Our understanding of mechanisms linking obesity to respiratory failure is partly limited by use of BMI as a measure of adiposity. While BMI is readily available and low-cost,^19^ it fails to capture differences in adipose tissue distribution, which vary by sex, race, and ethnicity.^20–24^ Adipose distribution is associated with both cardiovascular and all-cause mortality in the general population, independent of BMI.^20,25–27^ When energy intake exceeds energy utilization, excess energy is stored as free fatty acids, preferentially in subcutaneous adipose tissue depots.^28^ With continued energy excess, the storage capacity of subcutaneous depots can get overwhelmed and these free fatty acids are increasingly stored in ectopic locations (e.g. visceral adipose, pericardial adipose, and liver).^29^ Greater ectopic adiposity is associated with increased systemic inflammation, changes in innate immune cell function, and increased cardiovascular disease, kidney disease, insulin resistance, and diabetes mellitus.^30–33^

Whether greater ectopic fat storage alters risk of respiratory failure, such as with SARS-CoV-2 infection, is unknown. Prior work investigating the association between adiposity and outcomes from SARS-CoV-2 has relied on small single-center cohorts using clinically-indicated computed tomography (CT) imaging and is therefore limited by indication bias.^34–39^ Hence, we evaluated the association between adipose tissue depot sizes on pre-pandemic research CT scans and the risk of hospitalization or death from SARS-CoV-2 infection in the Collaborative Cohort of Cohorts for COVID-19 Research (C4R) study.^40^ We hypothesized that greater pericardial (PAT), abdominal visceral adipose tissue (VAT), and hepatic adipose deposition on pre-pandemic CT scans would associate with increased risk of hospitalization or death from SARS-CoV-2.

## Methods

C4R attempted to enroll all participants from 14 large US-based longitudinal cohorts who were alive on March 1, 2020 and had not withdrawn consent for cohort participation. Enrollment in C4R included completion of two serial questionnaires (April 2020 through February 2023) and ascertainment of COVID-related hospitalizations and deaths by questionnaires or active surveillance. Additional information on protocols and mechanisms of pooling clinical and demographic data across studies has been published.^40^ Four of the studies participating in C4R had available pre-pandemic CT-measures of adipose depots: the Multi-Ethnic Study of Atherosclerosis (MESA),^41^ the Jackson Heart Study (JHS),^42^ the Coronary Artery Risk Development in Young Adults (CARDIA) study,^43^ and the Framingham Heart Study (FHS).^44^ Descriptions of these cohorts are included in the *Supplementary Methods*.

We performed a retrospective analysis of C4R participants from the aforementioned cohorts who had any available CT-measure of adipose tissue prior to the pandemic. Cohorts obtained institutional review board approval from participating centers. Each cohort had previously consented participants to in-person, telephone, and/or e-mail contact and for abstraction of medical records. Consent for COVID-19 data collection was obtained in accordance with cohort-specific procedures. Participants with missing data for incident SARS-CoV- 2 infection were excluded from this report. Recurrent SARS-CoV-2 infections, which were rare, were not analyzed.

### Measurement of exposures

Methods for participant sampling for CT and protocols for adipose depot measurement in each cohort are further described in the *Supplementary Methods*. Chest and abdominal CT scans were performed as part of the research protocol in JHS from Exam 2 (2007-2010) and CARDIA Year 25 (2010-2011).^45–47^ Abdominal CT scans were performed as part of the research protocol in FHS Offspring, Generation 3, Omni 1, and Omni 2 cohorts (2008-2011). Chest CT and abdominal CT scans performed as part of the research protocol in MESA from Exams 1-4 (2000-2007) and Exams 2-3 (2002-2004), respectively.^47–49^ Pericardial adipose tissue (PAT) volume quantification protocols were identical in all three cohorts with available measures (CARDIA, JHS, MESA).

Primary analyses pooled PAT volumes across cohorts. The PAT distribution was skewed, therefore analyses were performed after log-2 transformation and results are reported per doubling of PAT volume. Measurement protocols for abdominal visceral adipose tissue (VAT), subcutaneous adipose tissue (SAT), and liver attenuation differed across all cohorts. FHS did not have available measures of liver attenuation. To pool these values, measures were standardized within each cohort and then the standardized values were combined for analyses; this approach was also used in secondary analyses of PAT.

### Measurement of outcomes

Our primary outcome was hospitalization or death from SARS-CoV-2 infection, hereafter described as “severe events.” This outcome was defined by self-report or active surveillance and confirmed, when available, by protocolized review of medical records or death certificates. Our secondary outcome was intensive care unit admission or death (hereafter, “critical events”) from SARS-CoV-2 infection, ascertained and confirmed in the same manner.

### Measurement of covariates

Covariates were obtained at the most recent pre-pandemic exam, which was not necessarily the time of CT (Supplemental Methods). BMI was calculated using height and weight, which were measured using standard methods at the most recent pre-pandemic exam. Vaccination date and status were obtained by self-report; vaccination status at the time of infection was obtained by comparing infection and vaccination dates.

Clinical cardiovascular disease was defined by self-reported physician diagnosis or adjudication/administrative criteria for myocardial infarction, angina pectoris, or stroke.

### Statistical Analysis

Adipose tissue measures were compared to BMI using Spearman correlations. For primary analyses, we used cause-specific hazards models to assess the association between adipose tissue measures and severe events over the period of March 1, 2020 to February 28, 2023. If a subject developed SARS-CoV-2 infection but did not experience a severe event, they were censored on the date of non-severe infection as subjects with prior non-severe infection are unlikely to be hospitalized or die of SARS-CoV-2 with a subsequent infection. Participants who had not developed SARS-CoV-2 infection were censored on last available day of follow-up, defined as the date of the most recently completed questionnaire. Models were stratified by cohort to account for potential differences in the baseline hazards function. We evaluated the proportional hazards assumption by regressing Schoenfeld residuals over time. We visually display the associations between adipose measures and outcomes using the “survival” package with the “pspline” function in R.^50,51^

We used directed acyclic graphs to identify a minimal set of covariates (Supplement Figure 1) including age, sex, race and ethnicity, educational attainment, smoking status, and vaccination status as a time-varying covariate. Among participants with missing vaccination data, vaccination status was classified as non-vaccinated as of March 1, 2021 (since vaccination was rare before that time point) and subsequent follow-up was censored. To evaluate independent effects of CT-measured adiposity, we adjusted for BMI, diabetes, hypertension, and cardiovascular disease in separate models. We evaluated subgroups defined a priori by sex, age at C4R enrollment (<65 or ≥65), smoking status (never, former, current), diabetes, cohort, race, and WHO BMI category.^52^ We evaluated for interactions using Likelihood Ratio tests.

We performed sensitivity analyses using Cox models with cohort as a frailty term using the “survival” package^51^ and Fine and Gray competing risks models stratified by cohort using the “crrSc” package.^53^ Frailty terms allow us to account for both between and within-study differences in risk.^54^ Fine and Gray models allow us to account for competing events without the assumption that these events are independent. For PAT, we performed additional sensitivity analyses with intra-cohort standardization. We used logistic regression models to evaluate the association between adipose depots and outcomes among participants with infection. These logistic regression models included cohort as a covariate.

All analyses were performed using R version 4.0.0 (R Foundation for statistical computing) on the C4R Analysis Commons.

## Results

Twelve thousand fifty-four participants from the 4 included cohorts were enrolled in C4R, of whom 8412 (70%) had at least one available CT-measure of any of the adipose depots of interest. Pericardial adipose measures were available from 6474 participants enrolled in MESA, CARDIA, and JHS from CT scans performed between the years 2000 and 2011 (Figure 1). VAT, SAT, and liver attenuation measures were available on 6467, 6323, and 7251 participants, respectively (Figure 1). Characteristics were similar across subgroups with different available adipose measures (Supplemental Table 1) and differed between cohorts by age, race and ethnicity, and education status, consistent with known differences in the populations enrolled for each study (Supplemental Methods, Supplemental Table 2). Participants with greater pericardial or visceral adiposity were older, more likely to be male, White, and with diabetes and/or cardiovascular disease, and less likely to be Black (Supplemental Tables 3-4).

**Figure 1:**
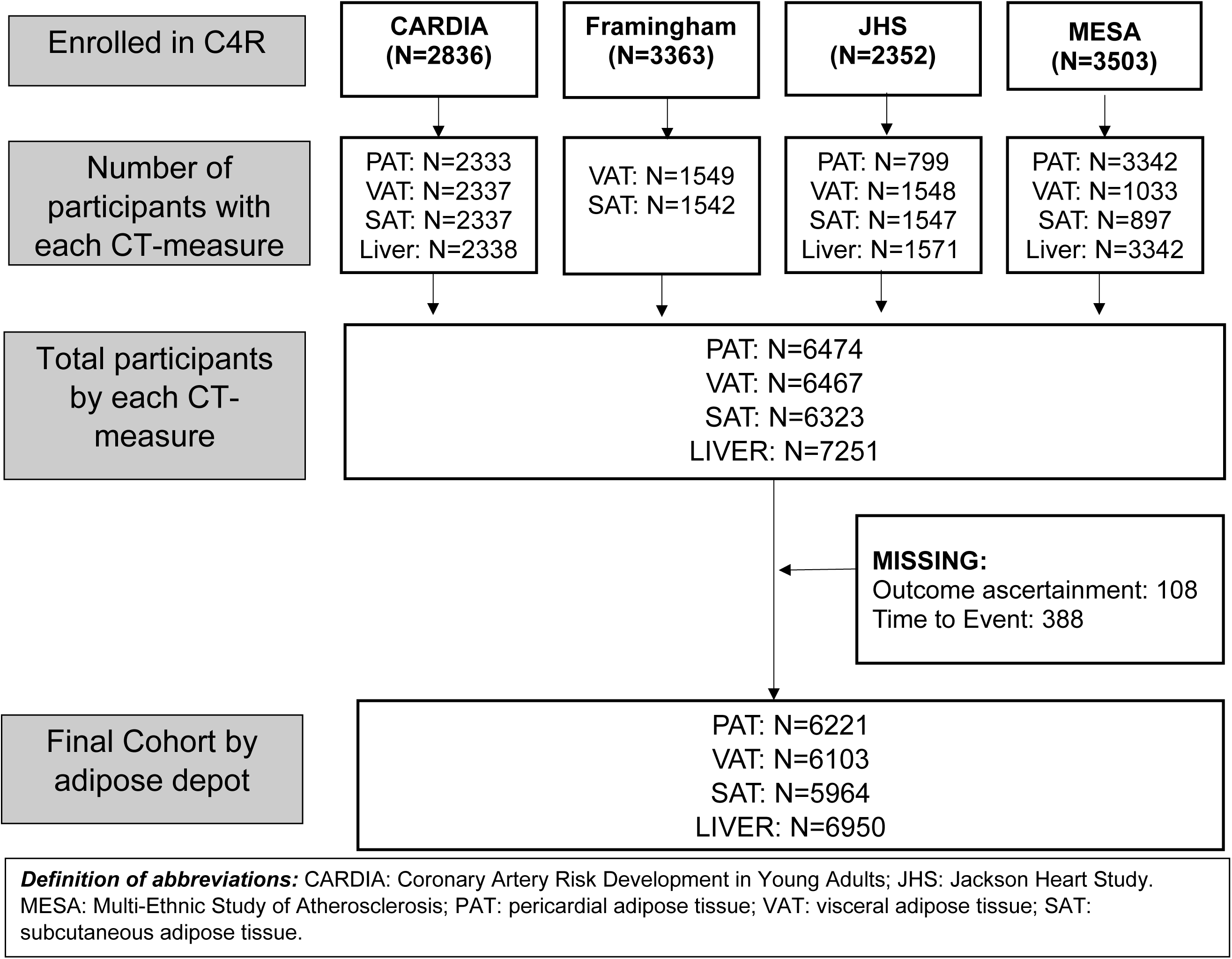
Flow Chart for study inclusion

One-hundred and eighty-four participants had a severe event and 794 had a non-severe infection over a median (interquartile range, IQR) follow-up of 547 (338-609) days. One-hundred thirty-nine of the severe events (76%) occurred within the first year of the pandemic when vaccine availability was low; only 8 severe events occurred among participants who had previously been vaccinated.

### BMI as a measure of adiposity

BMI was strongly correlated with SAT (r = 0.76, p<0.0001, Supplemental Table 5). BMI was only moderately correlated with VAT (r = 0.50, p<0.0001) and weakly correlated with PAT (r = 0.33, p<0.0001). Lower liver attenuation indicates greater adipose content. BMI was weakly inversely correlated with liver attenuation (r = -0.22, p<0.0001).

### Pericardial Adipose Tissue

In the cohort with PAT measured, 135 participants experienced a severe event over a median (IQR) of 365 (282-591) days. Greater PAT was associated with increased hazard of severe events in unadjusted and adjusted models (Table 1). In minimally-adjusted models, every doubling in PAT volume was associated with 64% increased hazards of severe event (95% CI 128-109%, p=0.0001, Figure 2A). This association remained after inclusion of BMI (HR 1.48, 95% CI 1.12-1.95), and diabetes, cardiovascular disease, and hypertension (HR 1.42, 95% CI 1.07-1.88). Greater pericardial adiposity was also associated with increased hazards of critical events in minimally-adjusted models (per doubling in PAT volume, HR 1.69, 95% CI 12-155%, p=0.013, Table 2). Findings were similar when PAT was standardized within cohorts and then analyzed (Supplemental Table 6), in analyses limited to those with documented infection (Supplemental Table 7), and in competing risk and frailty models (Supplemental Tables 7-8).

**Figure 2:**
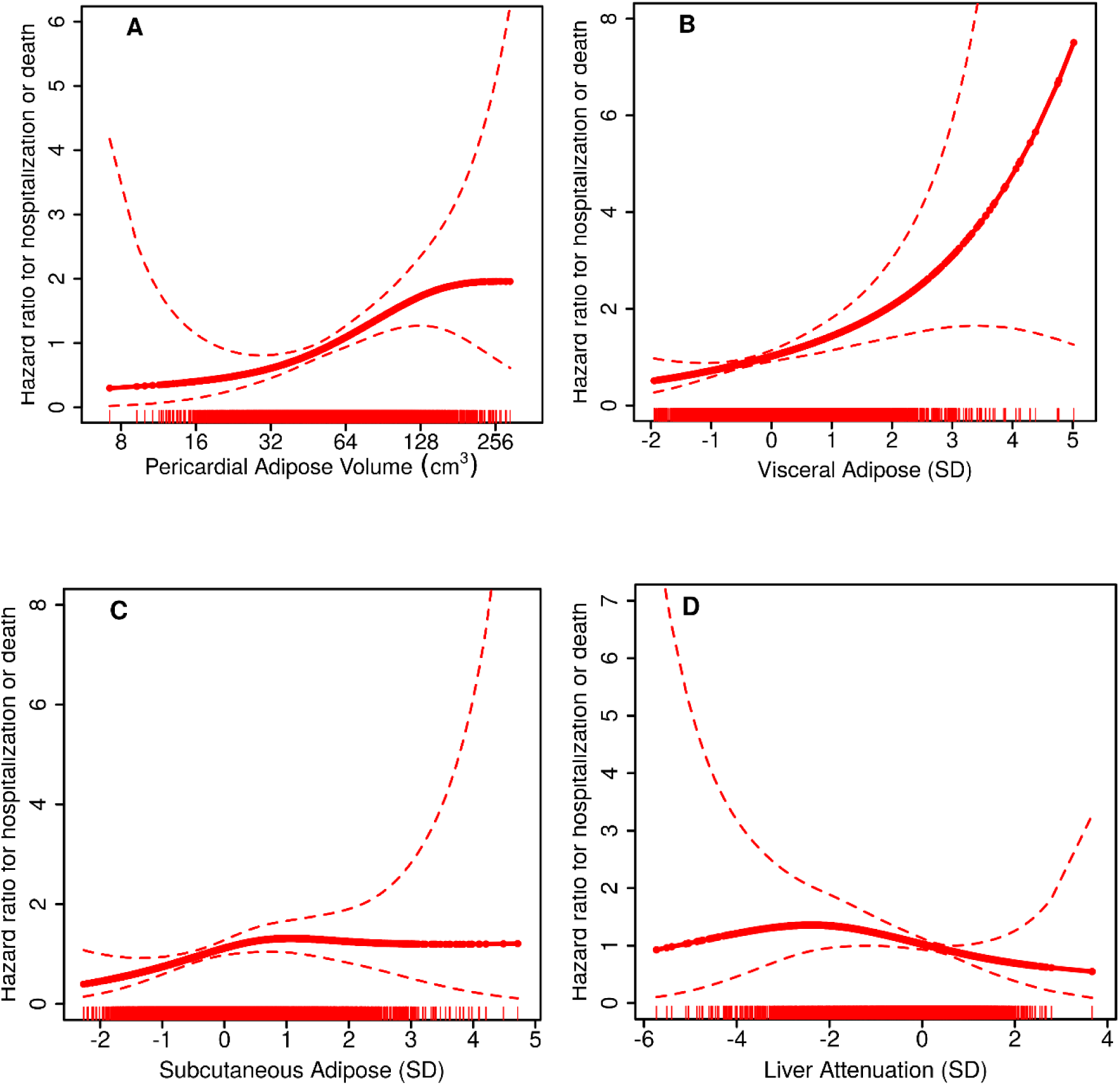
Continuous association between (A) pericardial adipose volume, (B) abdominal visceral adipose tissue, (C) abdominal subcutaneous adipose tissue, (D) liver attenuation (lower attenuation indicates greater lipid deposition) and hazard of hospitalization of death due to SARS-CoV2 infection. Models were adjusted for age, sex, race, education, smoking status, vaccination status at infection, and stratified by study. Point estimates for each subject are represented as individual dots (which appear as a continuous line) with dashed lines representing 95% confidence bounds. Each vertical line along the x-axis represents an individual subject.

**Table 1:**
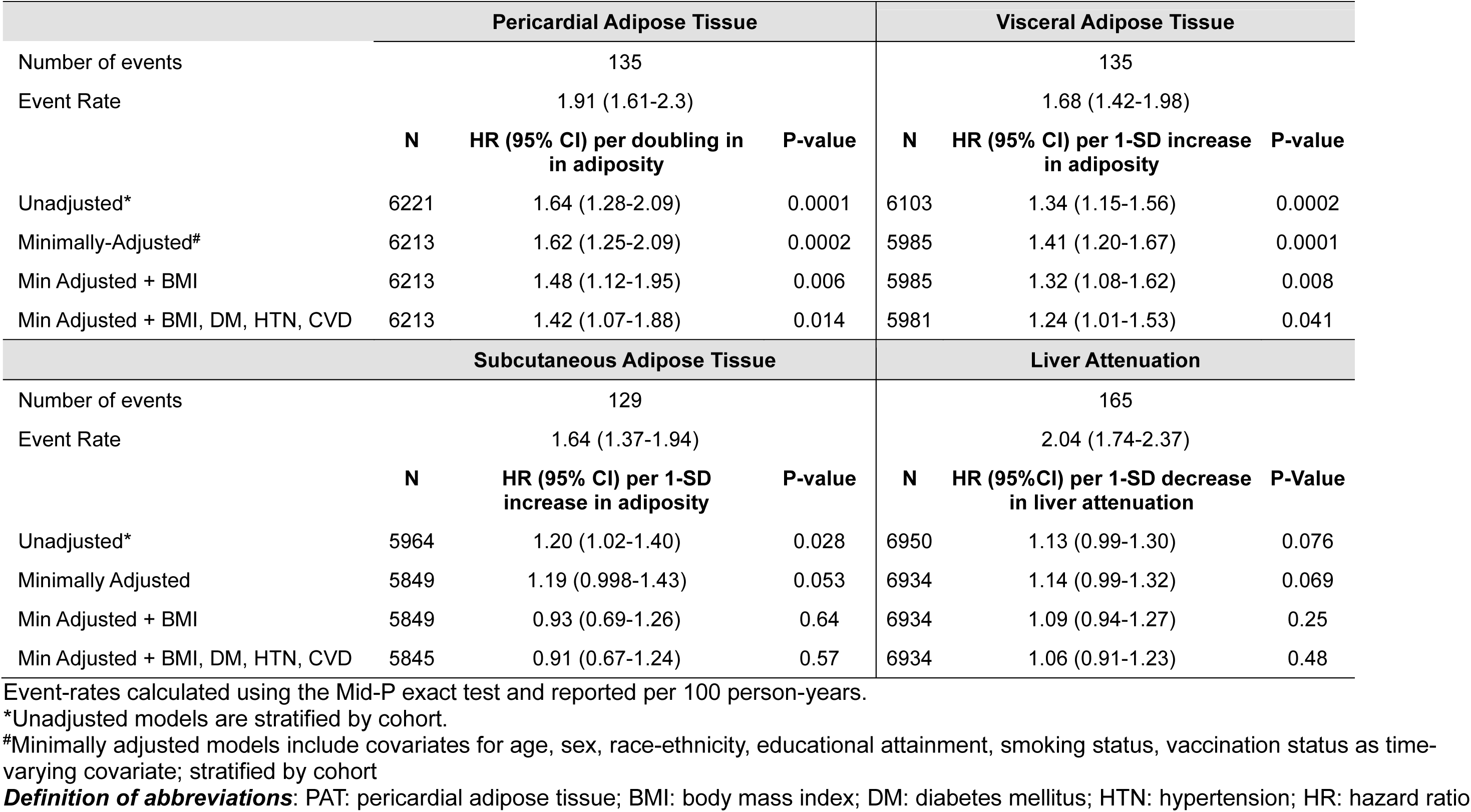
Association between adipose depot size and hazard of hospitalization or death due to SARS-CoV-2 infection.

**Table 2:**
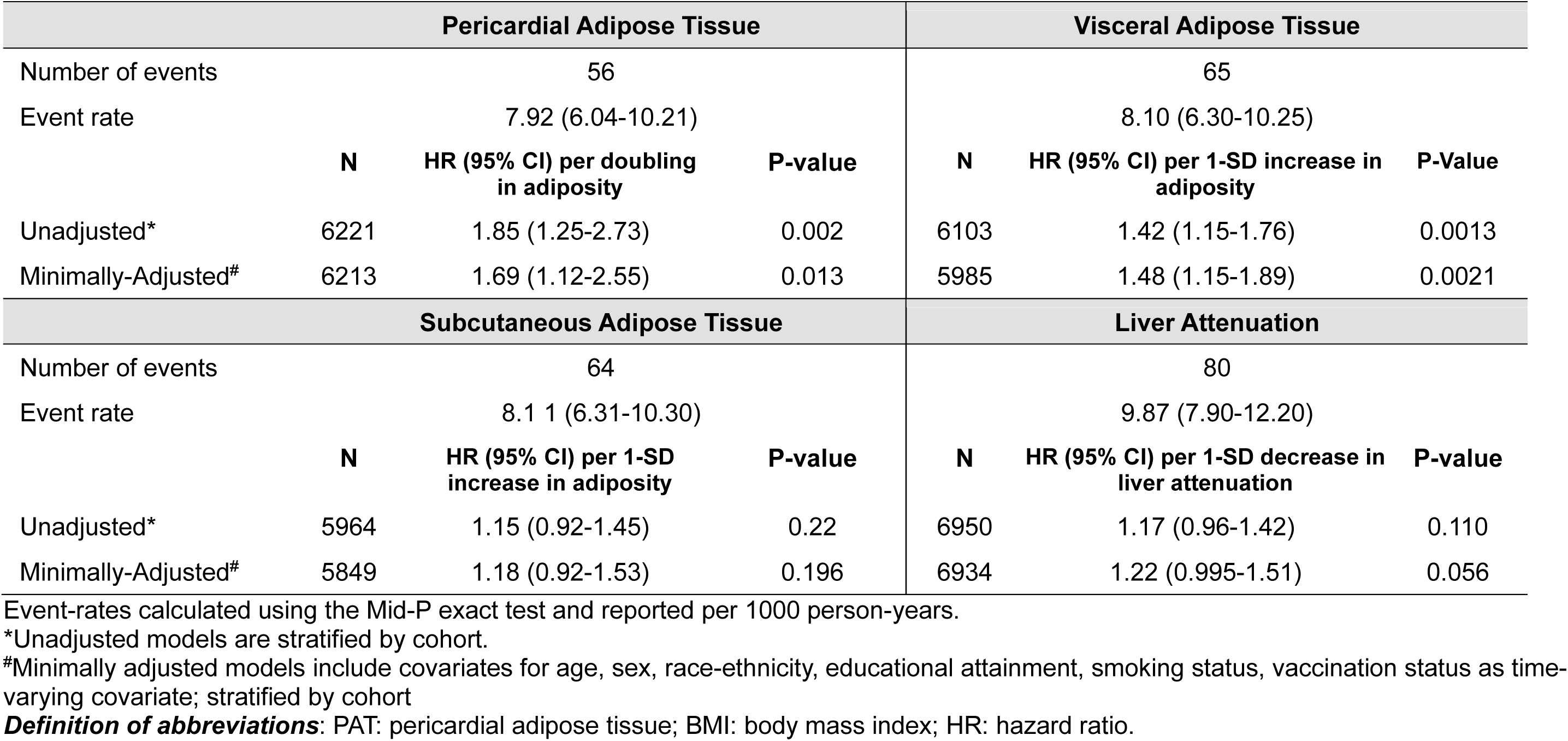
Association between adipose depot size and hazard of critical illness or death due to SARS-CoV-2 infection.

| Pericardial Adipose Tissue |  |  |  | Visceral Adipose Tissue |  |  |
| --- | --- | --- | --- | --- | --- | --- |
| Number of events | 56 |  |  | 65 |  |  |
| Event rate | 7.92 (6.04-10.21) |  |  | 8.10 (6.30-10.25) |  |  |
|  | **N** | **HR (95% CI) per doubling in adiposity** | **P-value** | **N** | **HR (95% CI) per 1-SD increase in adiposity** | **P-Value** |
| Unadjusted* | 6221 | 1.85 (1.25-2.73) | 0.002 | 6103 | 1.42 (1.15-1.76) | 0.0013 |
| Minimally-Adjusted<sup>#</sup> | 6213 | 1.69 (1.12-2.55) | 0.013 | 5985 | 1.48 (1.15-1.89) | 0.0021 |
| Subcutaneous Adipose Tissue |  |  |  | Liver Attenuation |  |  |
| Number of events | 64 |  |  | 80 |  |  |
| Event rate | 8.11 (6.31-10.30) |  |  | 9.87 (7.90-12.20) |  |  |
|  | **N** | **HR (95% CI) per 1-SD increase in adiposity** | **P-value** | **N** | **HR (95% CI) per 1-SD decrease in liver attenuation** | **P-value** |
| Unadjusted* | 5964 | 1.15 (0.92-1.45) | 0.22 | 6950 | 1.17 (0.96-1.42) | 0.110 |
| Minimally-Adjusted<sup>#</sup> | 5849 | 1.18 (0.92-1.53) | 0.196 | 6934 | 1.22 (0.995-1.51) | 0.056 |

Event-rates calculated using the Mid-P exact test and reported per 1000 person-years.
\*Unadjusted models are stratified by cohort.
<sup>#</sup>Minimally adjusted models include covariates for age, sex, race-ethnicity, educational attainment, smoking status, vaccination status as time-varying covariate; stratified by cohort
**Definition of abbreviations:** PAT: pericardial adipose tissue; BMI: body mass index; HR: hazard ratio.

### Abdominal visceral adipose

In the cohort with VAT measured, 135 participants experienced a severe event over a median (IQR) of 537 (365-610) days. Greater VAT was associated with increased hazards of severe events in unadjusted and adjusted models (Figure 2B, Table 1). In minimally adjusted models, every 1-SD increase in abdominal VAT was associated with 41% (95%CI 20-67%, p=0.0002) increased hazards of severe event. This association remained after inclusion of BMI (HR 1.32, 95% CI 1.09-1.56), and diabetes, cardiovascular disease, and hypertension (HR 1.24, 95% CI 1.01-1.53). Greater abdominal visceral adiposity was associated with increased hazards of critical events in minimally adjusted models (per 1-SD increase in VAT, HR 1.48, 95% CI 1.15-1.89, p=0.0021, Table 2). Findings were similar in sensitivity analyses (Supplemental Tables 7-9).

In models including both PAT and VAT, 1-SD increase in PAT was associated with 25% increased hazards (95% CI -4% to 63%) of severe events while 1-SD increase in VAT was associated with 23% increased hazards (95% CI -7% to 63%) in 3934 participants with available measures for both.

### Abdominal subcutaneous adipose

In the cohort with SAT measured, 129 participants experienced a severe event over a median (IQR) of 538 (365-610) days. Greater SAT was associated with increased hazards of hospitalization or death in unadjusted and minimally adjusted models (Table 2) but not after adjustment for BMI or obesity-related complications. The plot suggested that there may be a threshold effect wherein after a certain amount of SAT has accumulated, greater SAT no longer alters risk (Figure 2C). Associations were similar in sensitivity analyses (Supplemental Tables 8-9). SAT was not associated with risk of critical illness or death (Table 2), or with risk of hospitalization among those with infection after adjustment for confounders (Supplemental Table 9).

### Liver attenuation

In the cohort with hepatic adiposity measured, 165 participants experienced a severe event over a median (IQR) of 365 (308-426) days. Decreased liver attenuation (indicating more intra-hepatic adipose deposition) was not associated with significantly increased risk of severe events (HR 1.14, 95% CI 0.99-1.32, Table 1, Figure 2D) in primary analyses or sensitivity analyses (Supplemental Tables 7-8).

### Interactions

The associations between adipose depots and severe events were not modified by age, sex, smoking status, diabetes, cohort, race or BMI category (Figure 3, Supplemental Figure 2).

**Figure 3:**
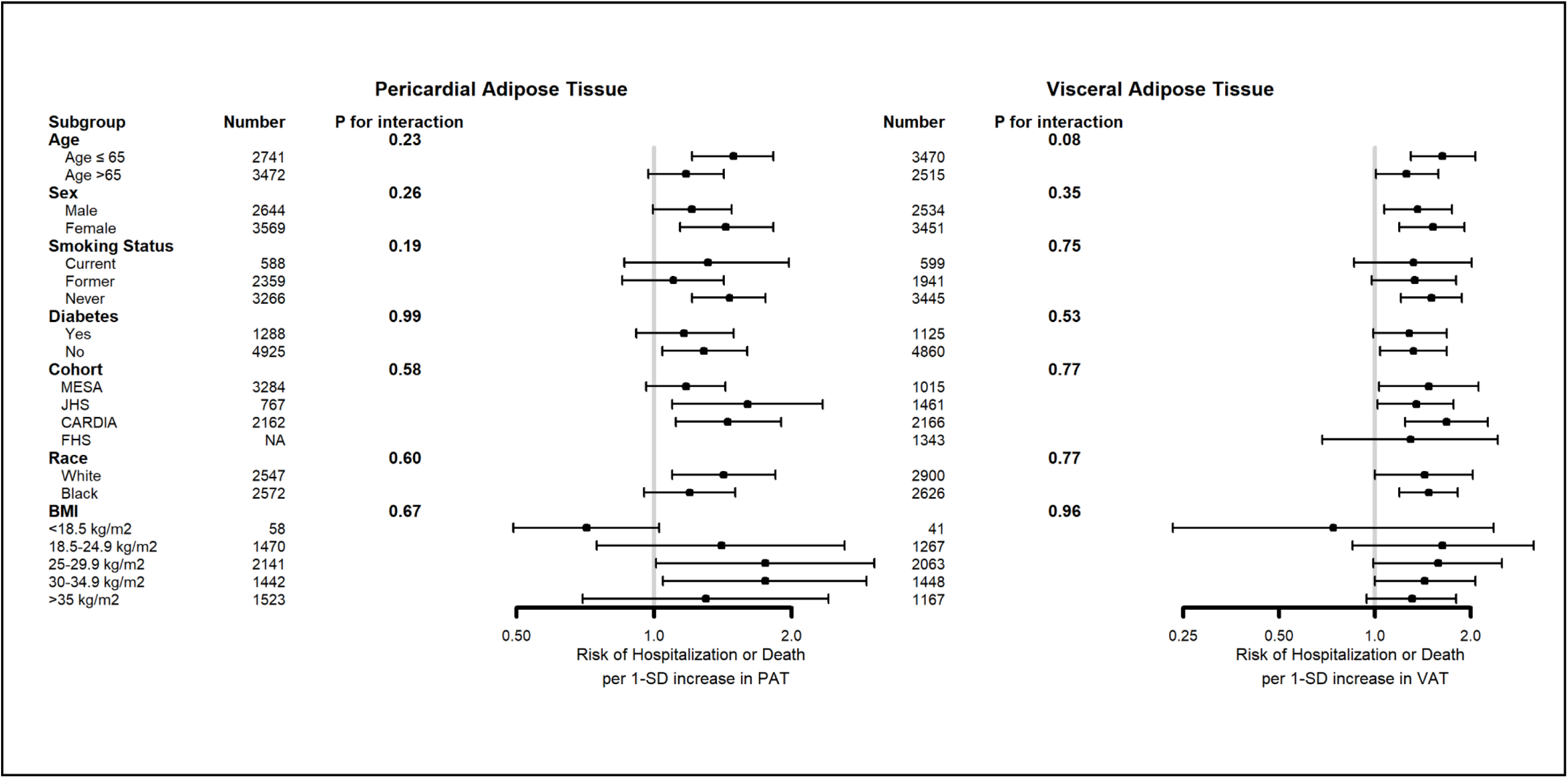
Forest plot demonstrating association between pericardial adipose, visceral adipose and hazard of hospitalization or death in subgroups defined by age, sex, smoking status, diabetes, cohort, and race. Models include same covariates as minimally-adjusted models (age, sex, race and ethnicity, educational attainment, smoking status, vaccination status as time-varying covariate; stratified by cohort) but exclude the variable of interest when evaluating subgroups defined by that variable.

## Discussion

Greater pericardial, abdominal visceral adipose, and abdominal subcutaneous adipose deposition, but not liver adipose deposition, as measured on pre-pandemic CT scans, were associated with increased risk of hospitalization or death from SARS-CoV-2 infection. Both pericardial and abdominal visceral adiposity, but not subcutaneous adiposity, were associated with risk of severe events even after adjustment for BMI and cardiometabolic comorbidities, suggesting an independent contribution of pericardial and abdominal visceral adipose to risk of hospitalization or death from SARS-CoV-2.

Our study is the first to evaluate pre-pandemic research CT-based measures of adiposity with risk of hospitalization or death due to SARS-CoV-2 infection. Prior work evaluating the association between CT measures of adiposity and outcomes from SARS-CoV-2 has had variable results.^34–39^ The differences are likely attributable to the selection bias inherent to single center retrospective studies of participants who required hospitalization and had a clinical indication for cross-sectional imaging. Use of pre-pandemic research cohorts with protocolized scans allowed us to evaluate the association between adiposity and disease severity on an unbiased, population level. We identified a consistent relationship between multiple measures of ectopic adipose deposition and risk of severe manifestations of SARS-CoV-2 infection. Our results are consistent with prior work demonstrating an association between research CT measures of adiposity and subclinical parenchymal lung injury in the general population,^55^ suggesting that there may be broader associations between adiposity and lung injury. Future work is needed to establish how CT-measured adiposity could be used to identify patients at high risk of respiratory failure in the setting of viral infection, such that they could be targeted for prevention and early treatment.

Our results illustrate that the standard, BMI-based classification of obesity does not capture all adiposity-related risk. Prior work has established that BMI frequently mis-classifies adiposity.^22,23,56,57^ Specifically, BMI has been shown to misclassify body fat status in 41% of subjects with heart failure and 70% of subjects with advanced lung disease.^56,57^ In our study, BMI did not strongly correlate with CT-measures of ectopic adipose deposition (PAT, VAT, liver attenuation), and associations of PAT and VAT with adverse

SARS-CoV-2 outcomes were only modestly attenuated by co-adjustment for BMI. Notably, associations of PAT and VAT with adverse outcomes were not attenuated by co-adjustment for diabetes, cardiovascular disease, and hypertension, which are known risk factors for severe SARS-CoV-2 infection.^58–60^ This suggests direct pathways linking adiposity and adverse SARS-CoV-2 outcomes.

Greater subcutaneous adiposity may increase risk of severe SARS-CoV-2 infection. Associations persisted after adjustment for a minimal set of confounders, but not after inclusion of BMI or cardiometabolic obesity complications. There may be a threshold effect between SAT and respiratory failure; greater SAT appeared to be associated with increased risk of respiratory failure until a threshold beyond which it no longer altered risk. Once subcutaneous adipose storage capacity is exceeded, fat begins to accumulate in other locations.^61^ Greater accumulation after this SAT storage capacity threshold has been reached, may not significantly alter its inflammatory state.

There are multiple potential mechanisms to explain why excess adiposity may predispose adults to severe manifestations of SARS-CoV-2 infection.^62^ Increased pro-inflammatory adipose tissue macrophages result in greater production of inflammatory mediators including interleukin-6, monocyte chemoattractant protein-1, and leptin.^63–68^ Pericardial adipose may particularly increase risk of respiratory failure as it drains directly into the coronary and pulmonary circulations; chronic release of these inflammatory mediators into the pulmonary circulation may lead to greater endothelial damage or alterations in immune cell function, that leave the lung primed for greater injury in the setting of subsequent infection.^32,66,69^ Greater expression of ACE-2 (the binding receptor SARS-CoV-2 virus) in visceral adipose depots could contribute to excess risk by creating a larger viral reservoir.^70^ Notably, obesity is a known risk factor for respiratory failure in multiple settings,^12,14,15,71–73^ suggesting that this association, and these mechanisms, may not be specific to the SARS-CoV-2 virus.

Future work should focus on whether these mechanisms could link ectopic adipose deposition to severe lung injury from viral infection and whether targeting ectopic adipose deposition could alter risk of lung injury.

There are limitations to our study. First, CT measures were obtained 10-20 years prior to the pandemic. Prior work has shown that these depots may increase over time.^74,75^. Future work should consider measuring pericardial and abdominal visceral adipose in cohorts with more recent imaging. Second, protocols for measurement of abdominal adipose tissue differed across cohorts. Reassuringly, associations were similar within cohorts and across two measures of adiposity. Future work should validate a single standard measurement protocol to increase the clinical utility of these measures. Third, ascertainment of mild infection is primarily dependent on testing and self-report. It is possible that some participants who we consider “at-risk” for incident infection were asymptomatically infected prior and should not have been part of the at-risk population. Fourth, approximately 6% of eligible cohort participants did not enroll in C4R, which may have introduced selection bias; however, reassuringly, characteristics of enrolled participants were similar to the eligible population.^40^

In conclusion, greater CT-measured pericardial and abdominal visceral adiposity were associated with increased risk of hospitalization or death from SARS-CoV-2 infection independent of BMI, diabetes, cardiovascular disease, and hypertension in a multi-ethnic US population-based sample of adults. Future work should investigate the use of CT measures of adipose to better quantify respiratory failure risk and to investigate mechanisms linking ectopic adipose deposition to risk of acute lung injury in the general population.

## Data Availability

Raw deidentified data is available, upon request and with appropriate consortium and cohort permissions, on the C4R Analysis Commons. C4R Analysis Commons, hosted on BioData Catalyst powered by Seven Bridges (https://accounts.sb.biodatacatalyst.nhlbi.nih.gov/).

https://c4r-nih.org

## Funding

NHLBI K23 150280, NHLBI K23 HL 141584 <u>The Collaborative Cohort of Cohorts for COVID-19 Research (C4R) Study</u> is supported by grant OT2HL156812 from the NHLBI Collaborating Network of Networks for Evaluating COVID-19 and Therapeutic Strategies/Researching COVID to Enhance Recovery, with co-funding from the National Institute of Neurological Disorders and Stroke (NINDS) and the National Institute on Aging (NIA).

<u>The Coronary Artery Risk Development in Young Adults (CARDIA) Study</u> is conducted and supported by the NHLBI in collaboration with the University of Alabama at Birmingham (contracts HHSN268201800005I and HHSN268201800007I), Northwestern University (contract HHSN268201800003I), the University of Minnesota (contract HHSN268201800006I), and the Kaiser Foundation Research Institute (contract HSN268201800004I).

<u>The Framingham Heart Study</u> has received support from the NHLBI (grant N01-HC-25195, contract HHSN268201500001I, and grant 75N92019D00031).

<u>The Jackson Heart Study</u> is supported by and conducted in collaboration with Jackson State University (contract HHSN268201800013I), Tougaloo College (contract HHSN268201800014I), the Mississippi State Department of Health (contract HHSN268201800015I), the University of Mississippi Medical Center (contracts HHSN268201800010I, HHSN268201800011I, and HHSN268201800012I), the NHLBI, and the National

Institute on Minority Health and Health Disparities.

<u>The Multi-Ethnic Study of Atherosclerosis (MESA)</u> and the MESA SNP Health Association Resource (SHARe) are conducted and supported by the NHLBI in collaboration with the MESA investigators. Support for MESA is provided by grants and contracts 75N92020D00001, HHSN268201500003I, N01-HC-95159, 75N92020D00005, N01-HC-95160, 75N92020D00002, N01-HC-95161, 75N92020D00003, N01-HC-95162, 75N92020D00006, N01-HC-95163, 75N92020D00004, N01-HC-95164, 75N92020D00007, N01-HC-95165, N01-HC-95166, N01-HC-95167, N01-HC-95168, N01-HC-95169, R01-HL077612, R01-HL093081, R01- HL130506, R01-HL127028, R01-HL127659, R01-HL098433, R01-HL101250, and R01-HL135009 from the NHLBI; grant R01-AG058969 from the NIA; and grants UL1-TR-000040, UL1-TR-001079, and UL1-TR-001420 from the National Center for Advancing Translational Sciences.

