## Supplemental for "Greater pericardial and visceral adiposity increase risk of hospitalization or death from SARS-CoV-2 infection in community dwelling adults: A C4R Study"

**Supplemental Methods 1:** Descriptions of included longitudinal cohort studies.

**Supplemental Methods 2:** Approach to adipose measurement for each cohort.

**Supplemental Methods 3:** Covariate measurements and definitions

**Supplemental Methods 4:** Description of method for participant sampling for each adipose measure by cohort

**Supplemental Figure 1:** Directed acyclic graph demonstrating absence of any paths between exposure and outcome after controlling for age, smoking status, socioeconomic status (educational attainment), race-ethnicity and cohort.

**Supplemental Figure 2:** Forest plot demonstrating association between subcutaneous adipose, liver attenuation and risk of hospitalization or death in subgroups defined by age, sex, smoking status, diabetes, cohort, and race.

**Supplemental Table 1:** Study subject characteristics for each available adipose measure

**Supplemental Table 2:** Study subject characteristics by cohort

**Supplemental Table 3:** Subject characteristics by quartile of pericardial adipose tissue volume ($cm^3$)

**Supplemental Table 4:** Subject characteristics by quartile of standardized visceral adipose tissue

**Supplemental Table 5:** Association between BMI and CT-measures of body composition

**Supplemental Table 6:** Association between one standard deviation increase in pericardial adipose tissue volume and outcomes

**Supplemental Table 7:** Association between adipose depot size and odds of hospitalization or death among those with documented infection

**Supplemental Table 8:** Association between adipose depot size and risk of hospitalization or death due to SARSCoV-2 infection in Competing Risks Models.

**Supplemental Table 9:** Association between adipose depot size and risk of hospitalization or death due to SARSCoV-2 infection in Cox Frailty models

**Supplemental References**

**Supplemental Methods 1:** Descriptions of included longitudinal cohort studies.

CARDIA

CARDIA is a study examining the development and determinants of clinical and subclinical CVD and their risk factors. It began in 1985-1986 with a cohort of 5115 Black and White men and women aged 18-30 years. The participants were selected so that there would be approximately the same number of people in subgroups of race (Black and White), gender (female and male), education (high school or less and more than high school) and age (18-24 and 25-30 years) in each of 4 field centers: Birmingham, AL; Chicago, IL; Minneapolis, MN; and Oakland, CA. These same participants were asked to participate in follow-up examinations during 1987-1988 (Year 2), 1990-1991 (Year 5), 1992-1993 (Year 7), 1995-1996 (Year 10), 2000-2001 (Year 15), 2005-2006 (Year 20), 2010-2011 (Year 25), 2015-2016 (Year 30), and 2020-2022 (Year 35). A majority of the group has been examined at each of the follow-up examinations (91%, 86%, 81%, 79%, 74%, 72%, 72%, 71%, and 67% [despite the impact of the COVID-19 pandemic on Year 35], respectively). While the specific aims of each examination have varied, data have been collected on a variety of factors believed to be related to heart disease. These include conditions with clear links to heart disease such as blood pressure, cholesterol and other lipids, and glucose. Data have also been collected on physical measurements such as weight and body composition as well as lifestyle factors such as dietary and exercise patterns, substance use (tobacco and alcohol), behavioral and psychological variables, medical and family history, and other chemistries (e.g., insulin). In addition, subclinical atherosclerosis has been measured via echocardiography during Years 5, 10, 25, and 30, a chest CT scan during Years 15, 20, 25, and 35, an abdominal CT scan during Years 25 and 35, and carotid ultrasound during Year 20. A brain MRI was performed on a subset of participants in Years 25, 30, and 35. The CARDIA cohort, born between 1955 and 1968, has been influenced by the obesity epidemic at ages younger than participants in other established NHLBI cohorts. Further investigation of the mechanisms linking obesity to derangements in cardiovascular structure and function and the etiology of clinical events promises to generate important new knowledge to inform health promotion and disease prevention efforts.[1]

Jackson Heart Study

The JHS is a community-based cohort study evaluating risk factors for cardiovascular and related diseases among adult African Americans residing in the three counties (Hinds, Madison, and Rankin) that make up the Jackson, Mississippi metropolitan area. Data and biologic materials have been collected from 5,306 participants, including a nested family cohort of 1,498 members of 264 families. The age at enrollment for the unrelated cohort was 35-84 years; the family cohort included related individuals >21 years old. Participants have provided extensive medical and psychosocial histories and had an array of physical and biochemical measurements and diagnostic procedures during a baseline examination (2000-2004) and two follow-up examinations (2005-2008 and 2009-2012). Samples for genomic DNA were collected during the first two examinations. Annual follow-up interviews and cohort surveillance of cardiovascular events and mortality are continuing, and a fourth examination is in progress.[2]

Framingham Heart Study

FHS was initiated in 1948. Researchers recruited 5,209 men and women between the ages of 30 and 62 from the town of Framingham, Massachusetts, and began the first round of extensive physical examinations and lifestyle interviews that they would later analyze for common patterns related to CVD development. Since 1948, the subjects have returned to the study every two years for an examination consisting of a detailed medical history, physical examination, and laboratory tests, and in 1971, the study enrolled a second-generation cohort – 5,124 of the original participants' adult children and their spouses – to participate in similar examinations. The second examination of the Offspring cohort occurred eight years after the first examination, and subsequent examinations have occurred approximately every four years thereafter. In April 2002, the Study entered a new phase: the enrollment of a third generation of participants, the grandchildren of the original cohort. The first examination of the Third Generation Study was completed in July 2005 and involved 4,095 participants. Thus, the FHS has evolved into a prospective, community-based, three generation family study. In addition to research studies focused on risk factors, subclinical CVD and clinically apparent CVD, Framingham investigators have also collaborated with leading researchers from around the country and throughout the world on projects involving some of the major chronic illnesses in men and women, including dementia, osteoporosis and arthritis, nutritional deficiencies, eye diseases, hearing disorders, and chronic obstructive lung disease.[3] [4]

MESA

MESA is a study of the characteristics of subclinical CVD (disease detected non-invasively before it has produced clinical signs and symptoms) and the risk factors that predict progression to clinically overt cardiovascular disease or progression of the subclinical disease.[5] MESA researchers study a diverse, population-based sample of 6,814 men and women aged 45-84 without known clinical cardiovascular disease. Thirty-eight percent of the recruited participants are White, 28 percent African American, 22 percent Hispanic, and 12 percent of Chinese descent. Participants were recruited from six field centers across the United States: Wake Forest University, Columbia University, Johns Hopkins University, University of Minnesota, Northwestern University and University of California – Los Angeles. At baseline, each participant received an extensive physical exam and determination of coronary artery calcification, ventricular mass and function, flow-mediated endothelial vasodilation, carotid intimal-medial wall thickness and presence of echogenic lucencies in the carotid artery, lower extremity vascular insufficiency, arterial wave forms, electrocardiographic (ECG) measures, standard coronary risk factors, sociodemographic factors, lifestyle factors, and psychosocial factors. Selected repetition of subclinical disease measures and risk factors at follow-up visits allows study of the progression of disease. Blood samples have been assayed for putative biochemical and genetic risk factors and stored for case-control studies. Participants are being followed for identification and characterization of cardiovascular disease events, including acute myocardial infarction and other forms of coronary heart disease (CHD), stroke, and heart failure; for CVD interventions; and for mortality. The first examination took place over two years, from July 2000 – July 2002. It has been followed by six examination periods that were 17-20 months in length. Participants have been contacted every 9 to 12 months throughout the study to assess clinical morbidity and mortality.

**Supplemental Methods 2:** Approach to adipose measurement for each cohort.

***Pericardial Adipose Tissue***

Jackson Heart, MESA, CARDIA

JHS scans used a 16-channel multidetector CT system with cardiac gating. MESA and CARDIA scans used either an ECG-triggered (80% of RR-interval) electron beam CT scanner or with prospectively ECG-triggered scan acquisition at 50% of RR interval with multidetector CT that acquired 4 simultaneous 2.5 mm slices for each cardiac cycle in a sequential or axial mode.[6 7] Pericardial fat volumes in the Jackson Heart study, MESA, and CARDIA were performed using the same validated protocol.[6-11] Eighteen slices, each 2.5 mm thick, starting 1.5 cm above and ending 3.0 cm below the superior extent of the left main coronary artery were selected resulting in 4.5 cm coverage along the head-foot axis. Adipose tissue was identified using a threshold of −190 to −30 Hounsfield Units. Volume accounted for both the number of pixels containing adipose tissue and the thickness of the slices. Measures had high reproducibility.

***Abdominal Visceral and Subcutaneous Adipose***

Jackson Heart Study

The research CT protocol used a 16-channel multidetector CT system. Quality control and image analysis were performed at a core reading center (Wake Forest University School of Medicine, Winston-Salem, NC).[12 13] Twenty-four contiguous 2-mm-thick slices centered on the lumbar disk space at L4–L5 were used to quantify VAT and SAT: 12 images before the center of the L4–L5 disk space and 12 images after the disk space. The abdominal muscular wall was first manually traced, and the fat volumes in different compartments were measured by semiautomatic segmentation technique. Volume analysis software (Advantage Windows; GE Healthcare, Waukesha, WI) used a threshold range of −190 to −30 Hounsfield units to identify voxels containing fat. The VAT and SAT volumes were the sum of VAT and SAT voxels over 24 slices. This approach had high inter-reader reproducibility of 0.95 for both VAT and SAT.

CARDIA

A non-contrast CT of the abdomen was performed with multidetector CT. Quality control was performed at a core reading center (Wake Forest University School of Medicine, Winston-Salem, NC). A 10-mm block of continuous slices (10 X 1 mm or 8 X 1.25 mm) centered on the lumbar disk space at L4–L5. Analysts segmented the images based on anatomic boundaries into the entire abdomen, abdominal wall, and intra-abdominal compartments using the Medical

Image Processing, Analysis, and Visualization application (http://mipav.cit.nih.gov/index.php). Fat was identified as voxels with an attenuation between −190 and −30 Hounsfield units. The interclass correlation coefficient for inter-reader comparisons was 0.98 for VAT, and intra- and inter-reader error were 2.4% and 6.7%, respectively, in 156 scans that were blinded and reevaluated.[14]

Framingham

Subjects underwent eight-slice multi-detector CT imaging of the abdomen (LightSpeed Ultra, General Electric, Milwaukee, WI).[15 16] A 125-mm block of 25 contiguous 5-mm thick slices were acquired above the level of S1. The abdominal muscular wall separating the visceral from the subcutaneous compartment was manually traced using Aquarius 3D Workstation (TeraRecon Inc. San Mateo, CA). Fat was identified as voxels with an attenuation of −195 to −45 Hounsfield Units. The inter-class correlation coefficients for inter-reader comparisons were 0.992 for VAT and 0.997 for SAT.  Similar high correlations were noted for intra-reader comparisons.

MESA

Abdominal CT scans were performed using electron beam CT or multi-detector CT scanners.[17] Six transverse slices from these scans were processed using MIPAV 4.1.2 software (provided by the NIH). The measure at L4–L5 was used in this analysis. Fat tissue was identified as being between -190 and -30 Hounsfield units.

***Liver Attenuation***

Jackson Heart Study

The multi-detector abdominal CT scans described above were used to measure liver attenuation.[13] The liver attenuation was measured in 3 regions of interest in the parenchyma of the right lobe of the liver, avoiding large vessels and any focal lesions. The mean hepatic attenuation (Hounsfield units) across the 3 regions of interest was calculated. The correlation coefficient between 2 different readers on a random selected sample of 60 participants was 0.98 for LA, indicating reliable reproducibility of CT imaging measurements.[18 19]

CARDIA

Abdominal CT scans described above were used to measure hepatic attenuation. Three circular regions of interest measuring 100 $mm^2$ were placed on 3 distinct CT slices at the level of the T12–L1 intervertebral space, avoiding vessels and any lesions.[14] The mean hepatic attenuation

(Hounsfield units) was calculated from these 9 total regions of interest. The interclass correlation coefficient between different readers on a random selected sample of 156 participants was 0.97 for LA, indicating high reproducibility of CT measured LA in this study.

MESA

Cardiac CT scans described above were used to measure liver attenuation. Three regions of interest (~1 $cm^2$ each) were measured in the right lobe of the liver, avoiding vessels and any lesions/cysts. The mean hepatic attenuation (Hounsfield Units) was calculated as the average density of three regions of interest.[20]

**Supplemental Methods 3:** Covariate measurements and definitions

Pre-pandemic exam data were collected using highly standardized research protocols that were similar or identical across cohorts, and all data were systematically harmonized. Age at enrollment in March 2020 was calculated from cohort enrollment age. Sex and educational attainment were self-reported. Race and ethnicity were self-reported according to fixed categories that differed by cohort, hence they were harmonized into a single classification of race and ethnicity (American Indian [AI], Asian, Black, Hispanic, Non-Hispanic White), following 2000 US Census definitions. [21] Smoking status was self-reported, with biochemical verification in a subset.[22] Ever smoking status was defined as at least 100 lifetime cigarettes, and current smoking status as smoking within the past 30 days. Diabetes was defined by self-report, fasting blood glucose ≥126 mg/dL, or the use of insulin or hypoglycemic medications. Hypertension was defined by self-report, systolic blood pressure ≥140 mmHg, diastolic blood pressure ≥90 mmHg, or use of antihypertensive medications.

**Supplemental Methods 4:** Description of method for participant sampling for each adipose measure by cohort

| Study | Sampling Method |
|---|---|
| **Pericardial Adipose Tissue** | |
| JHS | 4203 eligible: 1414 underwent CT for PAT at exam, random sample after excluding subjects with weight > 350 pounds, pregnant, female <40, male <35. |
| MESA | Measured on all 6814 participants at Exam 1, and subset of participants enrolled in MESA ancillary studies (Exam 5, N=3302; Exam 4, N=1405; Exam 3, N=2805; Exam 2, N=2955). Criteria for enrollment in ancillary studies were the same as in the parent MESA study.[10 23] |
| CARDIA | 5115 patients initially enrolled in CARDIA; 3189 people agreed to undergo CT at Y25 out of a total of 3498 Y25 participants.[24] |
| **Abdominal Adipose Tissue** | |
| JHS | 4203 eligible: 2884 underwent abdominal CT, random sample after excluding subjects with weight >350 pounds (n=41), pregnant (n=13), female <40 (n=128), male < 35 (n=48).[13] |
| MESA | Random subset of enrolled subjects underwent abdominal CT scan at Exam 2 or 3. Of these, a small number were re-scanned at Exam 4.[17] |
| CARDIA | 5115 patients initially enrolled in CARDIA; 3189 people agreed to undergo CT at Y25 out of a total of 3498 Y25 participants.[24] |
| FHS | Original study enrollment numbers were: Offspring (N=5124); Gen 3 (N=4095); OMNI 1 (N=507), OMNI 2 (N=410). FHS-MDCT2 included 3052 subjects – anyone enrolled in FHS-MDCT1 and all OMNI participants. Gen3 and OMNI 2 participants had to attend FHS Exam 2 and satisfy age requirements (male >35 and female >40). |
| **Liver Attenuation** | |
| JHS | 4203 eligible: 2884 underwent abdominal CT, random sample after excluding subjects with weight >350 pounds (n=41), pregnant (n=13), female <40 (n=128), male < 35 (n=48).[13] |
| MESA | Measured on all 6814 participants at Exam 1, and subset of participants enrolled in MESA ancillary studies (Exam 5, N=3302; Exam 4, N=1405; Exam 3, N=2805; Exam 2, N=2955). Criteria for enrollment in ancillary studies were the same as in the parent MESA study. |
| CARDIA | 5115 patients initially enrolled in CARDIA; 3189 people agreed to undergo CT at Y25 out of a total of 3498 Y25 participants.[24] |

**Supplemental Figure 1:** Directed acyclic graph demonstrating absence of any paths between exposure and outcome after controlling for age, smoking status, socioeconomic status (educational attainment), race-ethnicity and cohort.

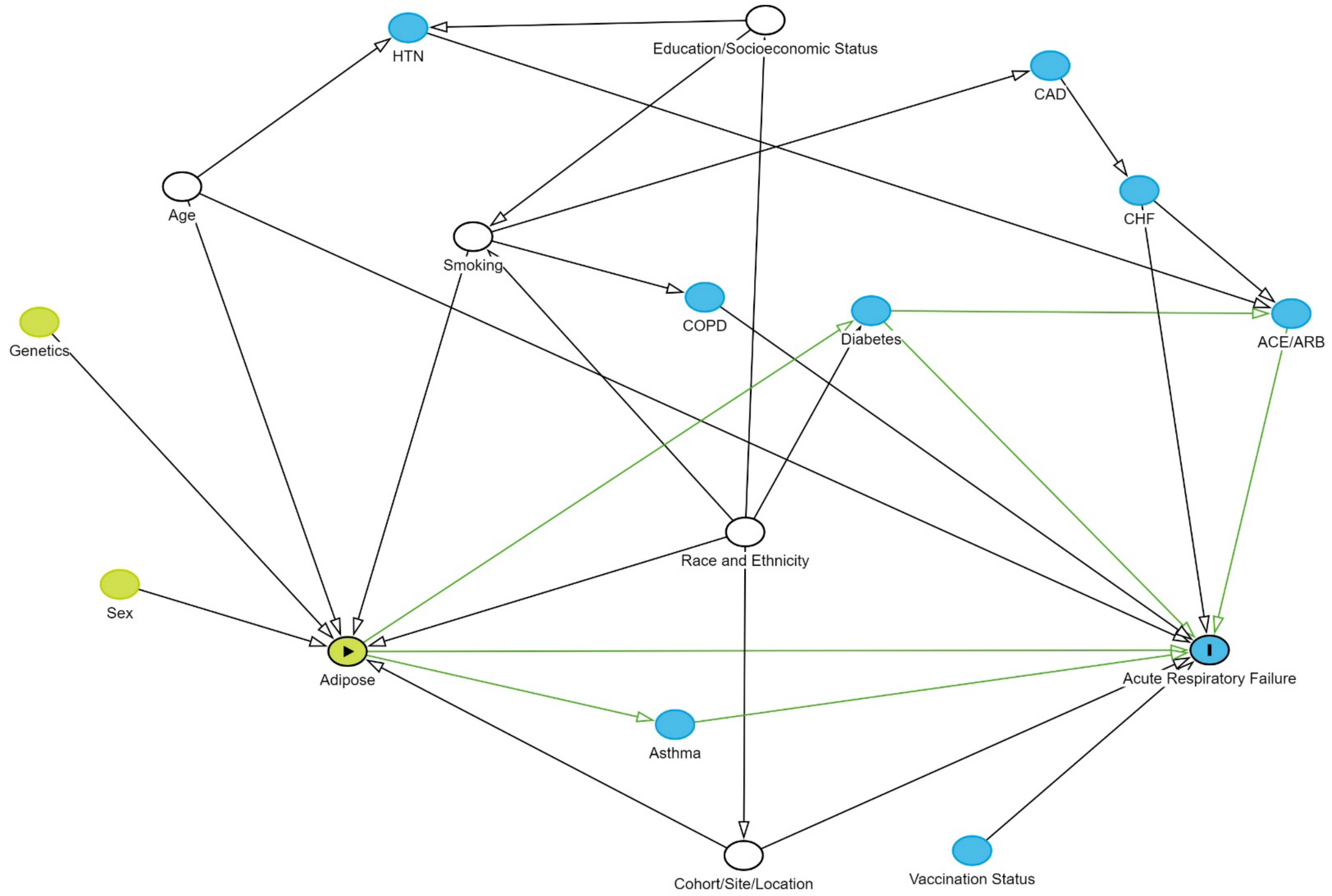


***Definition of Abbreviations***: HTN: hypertension; CAD: coronary artery disease; ACE: angiotensin converting enzyme inhibitor; ARB: angiotensin receptor blocker; CHF: congestive heart failure; COPD: chronic obstructive pulmonary disease.

**Supplemental Figure 2:** Forest plot demonstrating association between subcutaneous adipose, liver attenuation and hazard of hospitalization or death in subgroups defined by age, sex, smoking status, diabetes, cohort, and race. Models include same covariates as minimally-adjusted models (age, sex, race and ethnicity, educational attainment, smoking status, vaccination status as time-varying covariate; stratified by cohort) but exclude the variable of interest when evaluating subgroups defined by that variable.

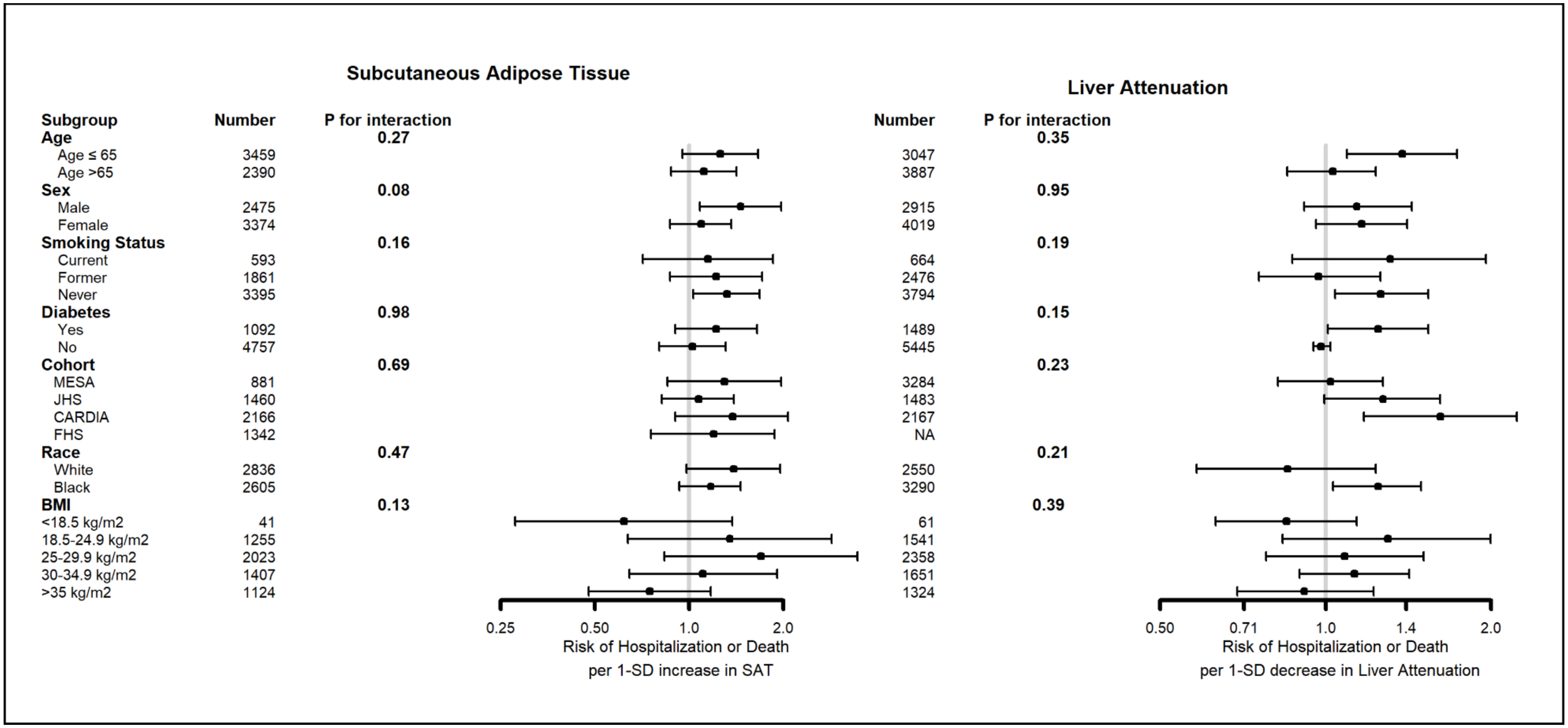

**Supplemental Table 1:** Study subject characteristics for each available adipose measure

| | Pericardial (N=6221) | Abdominal Visceral (N=6103) | Abdominal Subcutaneous (N=6964) | Liver Attenuation (N=7007) | Total (N=8412) |
|---|---|---|---|---|---|
| **Female, Sex** | 57% | 58% | 58% | 58% | 57% |
| **Age, years** | 69.8 (10.3) | 66.4 (9.4) | 66.2 (9.3) | 69.5 (10.1) | 69.1 (10.4) |
| **BMI, kg/m$^2$** | 29.7 (6.5) | 30.0 (6.5) | 30.0 (6.5) | 29.9 (6.5) | 29.7 (6.4) |
| **Race/Ethnicity** | | | | | |
| **White** | 41% | 49% | 49% | 37% | 46% |
| **Asian** | 7% | 3% | 3% | 6% | 6% |
| **Black** | 41% | 44% | 44% | 48% | 40% |
| **Hispanic** | 10% | 5% | 4% | 9% | 8% |
| **AIAN** | 0% | 0.1% | 0.2% | 0% | 0.1% |
| **Other** | 0% | 0.2% | 0.2% | 0% | 0.1% |
| **Education** | | | | | |
| **<High School** | 10% | 7% | 7% | 10% | 8% |
| **High School** | 20% | 18% | 18% | 20% | 18% |
| **Some College** | 27% | 22% | 22% | 25% | 24% |
| **College +** | 43% | 53% | 53% | 45% | 50% |
| **Smoking Status** | | | | | |
| **Never** | 53% | 57% | 58% | 55% | 55% |
| **Former** | 38% | 33% | 32% | 36% | 36% |
| **Current** | 9% | 10% | 10% | 10% | 9% |
| **Diabetes** | 21% | 19% | 19% | 21% | 20% |
| **Hypertension** | 61% | 56% | 55% | 62% | 59% |
| **Cardiovascular Disease** | 7% | 7% | 7% | 7% | 7% |
| **Vaccination** | 53% | 65% | 65% | 54% | 60% |

Continuous variables reported as mean (standard deviation). Categorical variables reported as percent. **Definition of abbreviations:** BMI: body mass index. **Missing**: Age (n=1), Education (n=123), Smoking Status (n=45), cardiovascular disease (n=54), vaccination status (n=2785).

**Supplemental Table 2:** Participant characteristics by cohort

| | CARDIA (N=2177) | Framingham (N=1451) | Jackson Heart (N=1494) | MESA (N=3290) | Total (N=8412) |
|---|---|---|---|---|---|
| **Female, Sex** | 60% | 55% | 64% | 54% | 57% |
| **Age, years** | 60.1 (3.6) | 67.5 (9.8) | 67.3 (8.4) | 76.6 (8.0) | 69.1 (10.4) |
| **BMI, kg/m$^2$** | 30.3 (7.1) | 28.8 (5.5) | 32.3 (6.5) | 28.6 (5.8) | 29.7 (6.4) |
| **Race/Ethnicity** | | | | | |
| **White** | 56% | 92% | 0% | 41% | 46% |
| **Asian** | 0% | 2% | 0% | 13% | 6% |
| **Black** | 44% | 2% | 100% | 26% | 40% |
| **Hispanic** | 0% | 2% | 0% | 20% | 8% |
| **AIAN** | 0% | 1% | 0% | 0% | 0.1% |
| **Other** | 0% | 1% | 0% | 0% | 0.1% |
| **Education** | | | | | |
| **<High School** | 6% | 1% | 8% | 13% | 8% |
| **High School** | 27% | 8% | 16% | 17% | 18% |
| **Some College** | 32% | 19% | 6% | 29% | 24% |
| **College +** | 34% | 72% | 70% | 42% | 50% |
| **Smoking Status** | | | | | |
| **Never** | 54% | 53% | 74% | 47% | 55% |
| **Former** | 31% | 40% | 16% | 47% | 36% |
| **Current** | 15% | 6% | 10% | 6% | 9% |
| **Diabetes** | 17% | 13% | 26% | 23% | 20% |
| **Hypertension** | 46% | 46% | 72% | 68% | 59% |
| **CV Disease** | 5% | 7% | 9% | 7% | 7% |
| **Vaccinated** | 60% | 90% | 57% | 48% | 60% |
| **Pericardial Adipose (cm$^3$)** | 55.3 (32.4) | N/A | 67.2 (29.1) | 76.3 (38.6) | 67.9 (36.7) |

*Continuous variables reported as mean (standard deviation). Categorical variables reported as percent.* **Definition of abbreviations:** BMI: body mass index.

**Supplemental Table 3:** Participant characteristics by quartile of pericardial adipose tissue volume ($cm^3$)

| | Lowest quartile (n=1556) | $2^{nd}$ quartile (n=1555) | $3^{rd}$ quartile (n=1555) | Highest quartile (n=1555) |
|---|---|---|---|---|
| **Female, Sex** | 72% | 61% | 55% | 42% |
| **Age, Mean (SD)** | 65.4 (9.2) | 69.6 (10.5) | 71.2 (10.3) | 74.1 (10.5) |
| **BMI, Mean (SD)** | 26.8 (5.5) | 29.3 (6.1) | 30.4 (6.4) | 32.0 (6.7) |
| **Race/Ethnicity** | | | | |
| **White** | 40% | 39% | 39% | 46% |
| **Asian** | 4% | 7% | 10% | 7% |
| **Black** | 52% | 45% | 39% | 30% |
| **Hispanic** | 5% | 9% | 12% | 17% |
| **AIAN** | 0% | 0% | 0% | 0% |
| **Other** | 0% | 0% | 0% | 0% |
| **Education** | | | | |
| **<High School** | 7% | 9% | 11% | 12% |
| **High School** | 22% | 20% | 18% | 21% |
| **Some College** | 27% | 29% | 28% | 25% |
| **College +** | 44% | 43% | 43% | 41% |
| **Smoking Status** | | | | |
| **Never** | 56% | 54% | 53% | 47% |
| **Former** | 32% | 38% | 38% | 44% |
| **Current** | 11% | 8% | 9% | 9% |
| **Diabetes** | 12% | 17% | 22% | 32% |
| **Hypertension** | 46% | 60% | 65% | 71% |
| **Cardiovascular Disease** | 4% | 6% | 6% | 10% |
| **Vaccination** | 56% | 53% | 53% | 50% |

*Continuous variables reported as mean (standard deviation). Categorical variables reported as percent.*
**Definition of abbreviations:** BMI: body mass index.

**Supplemental Table 4:** Participant characteristics by quartile of standardized visceral adipose tissue

| | Lowest quartile (N=1526) | $2^{nd}$ quartile (N=1526) | $3^{rd}$ quartile (N=1526) | Highest quartile (N=1525) |
|---|---|---|---|---|
| **Female, Sex** | 76% | 64% | 54% | 37% |
| **Age, Mean (SD)** | 65.0 (8.9) | 66.0 (9.2) | 67.0 (9.8) | 68.1 (9.6) |
| **BMI, Mean (SD)** | 25.5 (8.9) | 29.0 (5.3) | 31.6 (6.0) | 34.0 (6.6) |
| **Race/Ethnicity** | | | | |
| **White** | 50% | 43% | 46% | 57% |
| **Asian** | 4% | 4% | 3% | 1% |
| **Black** | 43% | 49% | 46% | 36% |
| **Hispanic** | 2% | 4% | 5% | 7% |
| **AIAN** | 0.1% | 0.2% | 0.2% | 0.1% |
| **Other** | 0.1% | 0.3% | 0.3% | 0.1% |
| **Education** | | | | |
| **<High School** | 5% | 7% | 7% | 9% |
| **High School** | 16% | 18% | 20% | 18% |
| **Some College** | 20% | 23% | 23% | 24% |
| **College +** | 59% | 52% | 50% | 49% |
| **Smoking Status** | | | | |
| **Never** | 60% | 61% | 57% | 51% |
| **Former** | 30% | 30% | 33% | 38% |
| **Current** | 9% | 9% | 10% | 11% |
| **Diabetes** | 7% | 14% | 23% | 32% |
| **Hypertension** | 39% | 52% | 62% | 69% |
| **Cardiovascular Disease** | 4% | 6% | 9% | 9% |
| **Vaccination** | 66% | 63% | 66% | 64% |

*Continuous variables reported as mean (standard deviation). Categorical variables reported as percent*
**Definition of abbreviations:** BMI: body mass index.

**Supplemental Table 5:** Association between BMI and CT-measures of body composition

| | Body Mass Index | Pericardial adipose | Visceral adipose | Subcutaneous adipose | Liver Attenuation |
|---|---|---|---|---|---|
| **Body Mass Index** | 1 | | | | |
| **Pericardial adipose** | 0.33* | 1 | | | |
| **Visceral adipose** | 0.50* | 0.74* | 1 | | |
| **Subcutaneous adipose** | 0.76* | 0.29* | 0.38* | 1 | |
| **Liver Attenuation$** | -0.22* | -0.32* | -0.44* | -0.19* | 1 |

*p<0.0001

$Lower liver attenuation indicates greater lipid content.

***Definition of abbreviations***: BMI: body mass index.

**Supplemental Table 6:** Association between one standard deviation increase in pericardial adipose tissue volume and outcomes

| | N | HR (95%CI) | P-value |
|---|---|---|---|
| **Risk of Hospitalization or Death** | | | |
| Unadjusted* | 6221 | 1.30 (1.13-1.50) | 0.0002 |
| Minimally-Adjusted# | 6213 | 1.29 (1.12-1.50) | 0.0007 |
| Min Adjusted + BMI | 6213 | 1.22 (1.03-1.43) | 0.018 |
| Min Adjusted + BMI, DM, HTN, CVD | 6213 | 1.19 (1.01-1.40) | 0.042 |
| **Risk of Critical Illness or Death** | | | |
| Unadjusted* | 6221 | 1.40 (1.14-1.73) | 0.0015 |
| Minimally-Adjusted# | 6213 | 1.34 (1.06-1.70) | 0.013 |
| **Odds of Hospitalization or Death among infected** | | **OR (95% CI)** | **P-value** |
| Unadjusted* | 731 | 1.40 (1.14-1.73) | 0.0015 |
| Minimally-Adjusted# | 730 | 1.31 (1.05-1.62) | 0.015 |
| Min Adjusted + Alternative Vax Value | 558 | 1.40 (1.07-1.83) | 0.015 |

[#]Minimally adjusted models include covariates for age, sex, race-ethnicity, educational attainment, smoking status, vaccination status as time-varying covariate; stratified by cohort

***Definition of abbreviations***: PAT: pericardial adipose tissue; BMI: body mass index; HR: hazard ratio

**Supplemental Table 7:** Association between adipose depot size and odds of hospitalization or death among those with documented infection

| | Pericardial Adipose Tissue | | | Visceral Adipose Tissue | | |
|---|---|---|---|---|---|---|
| | **N** | **OR (95% CI) per doubling in adiposity** | **P-value** | **N** | **OR (95% CI) per 1-SD increase in adiposity** | **P-value** |
| Unadjusted | 731 | 1.75 (1.36-2.25) | <0.0001 | 761 | 1.33 (1.10-1.59) | 0.003 |
| Minimally-Adjusted[#] | 730 | 1.59 (1.13-2.20) | 0.0003 | 746 | 1.46 (1.14-1.87) | 0.002 |
| Sensitivity Analysis[$] | 558 | 1.81 (1.15-2.85) | 0.0099 | 610 | 1.38 (0.98-1.94) | 0.063 |
| | **Subcutaneous Adipose Tissue** | | | **Liver Attenuation** | | |
| | **N** | **HR (95% CI) per 1-SD increase in adiposity** | **P-value** | **N** | **OR (95%CI) per 1-SD decrease in liver attenuation** | **P-Value** |
| Unadjusted | 736 | 1.35 (1.10-1.59) | 0.003 | 794 | 1.15 (0.97-1.35) | 0.108 |
| Minimally-Adjusted[#] | 721 | 1.14 (0.89-1.45) | 0.30 | 791 | 1.13 (0.93-1.39) | 0.22 |
| Sensitivity Analysis [$] | 592 | 1.01 (0.73-1.41) | 0.94 | 597 | 1.31 (0.996-1.72) | 0.054 |

[#]Minimally adjusted model include covariates for age, sex, race-ethnicity, educational attainment, smoking status, cohort, and vaccination status at time of infection if known and assumed to be unvaccinated if unknown
[$]Sensitivity Analysis includes vaccination status during follow-up assuming that those who were ever vaccinated, had been vaccinated at the time of infection and excluding those with unknown vaccination status.
***Definition of abbreviations***: PAT: pericardial adipose tissue; OR: odds ratio

**Supplemental Table 8:** Association between adipose depot size and risk of hospitalization or death due to SARSCoV-2 infection in Competing Risks Models.

| | Pericardial Adipose Tissue | | | Visceral Adipose Tissue | | |
|---|---|---|---|---|---|---|
| | **N** | **Sub-HR (95% CI) per doubling in adiposity** | **P-value** | **N** | **Sub-HR (95% CI) per 1-SD increase in adiposity** | **P-value** |
| Unadjusted* | 6221 | 1.62 (1.28-2.06) | <0.0001 | 6103 | 1.34 (1.16-1.56) | 0.0001 |
| Minimally-Adjusted[#] | 6213 | 1.53 (1.18-1.99) | 0.0012 | 5985 | 1.34 (1.12-1.60) | 0.001 |
| Min Adjusted + BMI | 6213 | 1.38 (1.04-1.82) | 0.024 | 5985 | 1.24 (0.99-1.54) | 0.058 |
| 0.60 | 6213 | 1.32 (1.01-1.74) | 0.041 | 5981 | 1.16 (0.93-1.45) | 0.18 |
| | **Subcutaneous Adipose Tissue** | | | **Liver Attenuation** | | |
| | **N** | **Sub-HR (95% CI) per 1-SD increase in adiposity** | **P-value** | **N** | **Sub-HR (95%CI) per 1-SD decrease in liver attenuation** | **P-Value** |
| Unadjusted* | 5964 | 1.19 (1.03-1.38) | 0.018 | 6950 | 1.13 (0.997-1.29) | 0.056 |
| Minimally Adjusted | 5849 | 1.19 (1.01-1.42) | 0.043 | 6934 | 1.12 (0.98-1.28) | 0.10 |
| Min Adjusted + BMI | 5849 | 0.95 (0.68-1.34) | 0.79 | 6934 | 1.07 (0.93-1.24) | 0.33 |
| Min Adjusted + BMI, DM, HTN, CVD | 5845 | 0.93 (0.66-1.31) | 0.68 | 6934 | 1.04 (0.90-1.19) | 0.60 |

*Unadjusted models are stratified by cohort.

[#]Minimally adjusted models include covariates for age, sex, race-ethnicity, educational attainment, smoking status, vaccination status as time-varying covariate; stratified by cohort

***Definition of abbreviations***: PAT: pericardial adipose tissue; BMI: body mass index; DM: diabetes mellitus; HTN: hypertension; HR: hazard ratio

**Supplemental Table 9:** Association between adipose depot size and risk of hospitalization or death due to SARSCoV-2 infection in Cox Frailty models

| | Pericardial Adipose Tissue | | | Visceral Adipose Tissue | | |
|---|---|---|---|---|---|---|
| | **N** | **HR (95% CI) per doubling in adiposity** | **P-value** | **N** | **HR (95% CI) per 1-SD increase in adiposity** | **P-value** |
| Unadjusted* | 6221 | 1.66 (1.30-2.11) | <0.0001 | 6103 | 1.35 (1.15-1.57) | 0.0002 |
| Minimally-Adjusted[#] | 6213 | 1.63 (1.26-2.10) | 0.0002 | 5985 | 1.41 (1.20-1.67) | 0.00005 |
| Min Adjusted + BMI | 6213 | 1.49 (1.13-1.96) | 0.005 | 5985 | 1.32 (1.08-1.61) | 0.008 |
| Min Adjusted + BMI, DM, HTN, CVD | 6213 | 1.43 (1.08-1.89) | 0.012 | 5981 | 1.25 (1.01-1.54) | 0.037 |
| | **Subcutaneous Adipose Tissue** | | | **Liver Attenuation** | | |
| | **N** | **HR (95% CI) per 1-SD increase in adiposity** | **P-value** | **N** | **HR (95%CI) per 1-SD decrease in liver attenuation** | **P-Value** |
| Unadjusted* | 5964 | 1.20 (1.02-1.40) | 0.0028 | 6950 | 1.14 (0.989-1.30) | 0.072 |
| Minimally Adjusted | 5849 | 1.20 (1.005-1.44) | 0.045 | 6934 | 1.14 (0.989-1.32) | 0.070 |
| Min Adjusted + BMI | 5849 | 0.95 (0.70-1.28) | 0.73 | 6934 | 1.09 (0.94-1.27) | 0.25 |
| Min Adjusted + BMI, DM, HTN, CVD | 5845 | 0.93 (0.68-1.26) | 0.63 | 6934 | 1.06 (0.91-1.23) | 0.48 |

*Unadjusted models are stratified by cohort.

[#]Minimally adjusted models include covariates for age, sex, race-ethnicity, educational attainment, smoking status, vaccination status as time-varying covariate; stratified by cohort

***Definition of abbreviations***: PAT: pericardial adipose tissue; BMI: body mass index; DM: diabetes mellitus; HTN: hypertension; HR: hazard ratio
